# The long COVID evidence gap: comparing self-reporting and clinical coding of long COVID using longitudinal study data linked to healthcare records

**DOI:** 10.1101/2023.02.10.23285717

**Authors:** Anika Knuppel, Andy Boyd, John Macleod, Nishi Chaturvedi, Dylan M Williams

**Affiliations:** MRC Unit of Lifelong Health and Ageing at UCL, University College London, London W1E 7HB, London, United Kingdom; Institute of Population Health Sciences, Bristol Medical School, University of Bristol, Bristol BS8 2BN, United Kingdom; NIHR Applied Research Collaboration (ARC) West, Bristol, UK

**Keywords:** COVID-19, long COVID, post-COVID syndrome, post-acute sequelae of COVID-19, UK LLC

## Abstract

The term “long COVID” (LC) was coined in spring 2020 by individuals with ongoing symptoms following COVID-19, but it took until December 2020 for clinical codes to be created in order to record persistent post-COVID-19 illness and referrals within electronic health records (EHRs). Analysis of whole-population EHR databases have helped understand the epidemiology of LC; yet concerns exist about the completeness of accessible EHRs for LC. UK longitudinal population studies (LPS) collected self-reported data on COVID-19 and LC from early 2020 and deposited these data in the UK Longitudinal Linkage Collaboration (UK LLC) research database where they are systematically linked to the participants EHRs. Comparisons of LPS reported LC with recorded LC in the EHRs of the same individuals may be helpful in understanding the epidemiology of emerging conditions such as LC. We used data from 10 UK LPS in the UK LLC to investigate whether participants self-reporting LC had a LC diagnosis or referral code in their English EHR after 10 to 22 months of follow up. Of 6412 participants with COVID-19 symptom duration data and linkage to health records, 898 (14.0%) self-reported LC of any severity in LPS surveys. Among these, just 42 (4.7%; 95% CI: 3.5, 6.3) were identified with LC-related codes in EHRs. In individuals reporting debilitating LC, this proportion was only marginally higher (5.6%; 95% CI: 3.7, 8.3). Our data show a striking discrepancy between LC as perceived and reported by participants in LPS and evidence of LC recorded in their EHRs; and that this discrepancy was patterned by ethnicity and possibly by indicators of deprivation. Self-reported symptoms may not be reflected in coded EHRs due to factors including variations in individuals help seeking behaviours, clinician coding practices and the availability of appropriate codes. However, these considerations appear unlikely to provide a complete explanation for the substantial observed reporting discrepancy. These results may indicate substantial unmet clinical need, in keeping with patient reports of difficulties accessing healthcare and sub-optimal recognition of, and response to, their illness when they do. They may also indicate potential shortcomings of epidemiological research on LC based on EHR- or LPS-based ascertainment alone and illustrate the value of triangulation between LPS and EHR data where linked and made available through resources such as the UK LLC.

## Introduction

The term ‘long COVID’ (LC) was coined in spring 2020 by individuals with ongoing symptoms following COVID-19, in response to unsatisfactory recognition of this emerging syndrome by healthcare practitioners. ^1^ In December 2020, clinical codes for persistent post-COVID-19 illness and related referrals were introduced and became available for use by practitioners to record details of clinical encounters in electronic health records (EHRs). EHRs, which have near whole population coverage, are increasingly used to help understand the epidemiology of disease alongside the effectiveness and safety of intervention. Many factors influence the completeness of information in EHRs including help-seeking behaviour of patients and the beliefs and data-recording behaviour of practitioners. Longitudinal population-based studies (LPS) often include participant reports of illness. These may be subject to reporting and participation biases. Comparing reported illness in studies to recorded illness in the EHRs of the same individuals may be helpful in understanding the epidemiology of emerging conditions such as LC.

## Methods

We investigated whether individuals with self-reported LC in 2020-21 had received an LC diagnosis or referral in English healthcare after 10 to 22 months of follow-up, among 6412 participants of ten LPS with COVID-19 survey data linked to their EHRs in the <u>UK Longitudinal Linkage Collaboration</u> (Supplementary material)

Self-reported LC was defined as reporting of four or more weeks of ongoing symptoms attributable to COVID-19, as per <u>National Institute for Health and Care Excellence</u> (NICE) guidelines. Seven LPS samples sought reports of debilitating symptoms only, with reports of any symptoms being sought in the remaining three LPS samples, as described in our previous research and the supplementary material. ^2^ LC-related healthcare interactions were identified from ICD-10 and SNOMED-CT codes (listed in the supplementary material) up to August 2022 from ‘Hospital Episodes Statistics’ (HES, the national database of English secondary care records) and ‘General Practice Extraction Service Data for Pandemic Planning and Research’ (<u>GDPPR, the national dataset of English COVID-19-relevant primary care records</u>), respectively.

## Results

Of 6412 participants with COVID-19 symptom duration data and linkage to health records, 898 (14.0%) self-reported LC of any severity in LPS surveys (Table S1, supplementary material). Among these, just 42 (4.7%; 95% CI: 3.5, 6.3) were identified with LC-related codes in EHR. When restricting to individuals reporting a history of debilitating LC, this proportion was only marginally higher (5.6%; 95% CI: 3.7, 8.3). Codes were assigned within a mean 3.8 and 4.9 months of symptom duration reporting, respectively. In analyses of differences in coding by sociodemographic characteristics (Table 1), likelihood of receiving an LC code differed by age – being highest in middle age and lower in younger and older LPS participants. Coding likelihood did not differ by sex. Those of white ethnicity were more likely to receive a code than individuals of other ethnicities. There was weak evidence that individuals of higher socioeconomic position were more likely to have EHR evidence of LC (*P* for trend = 0.18).

**Table 1:**
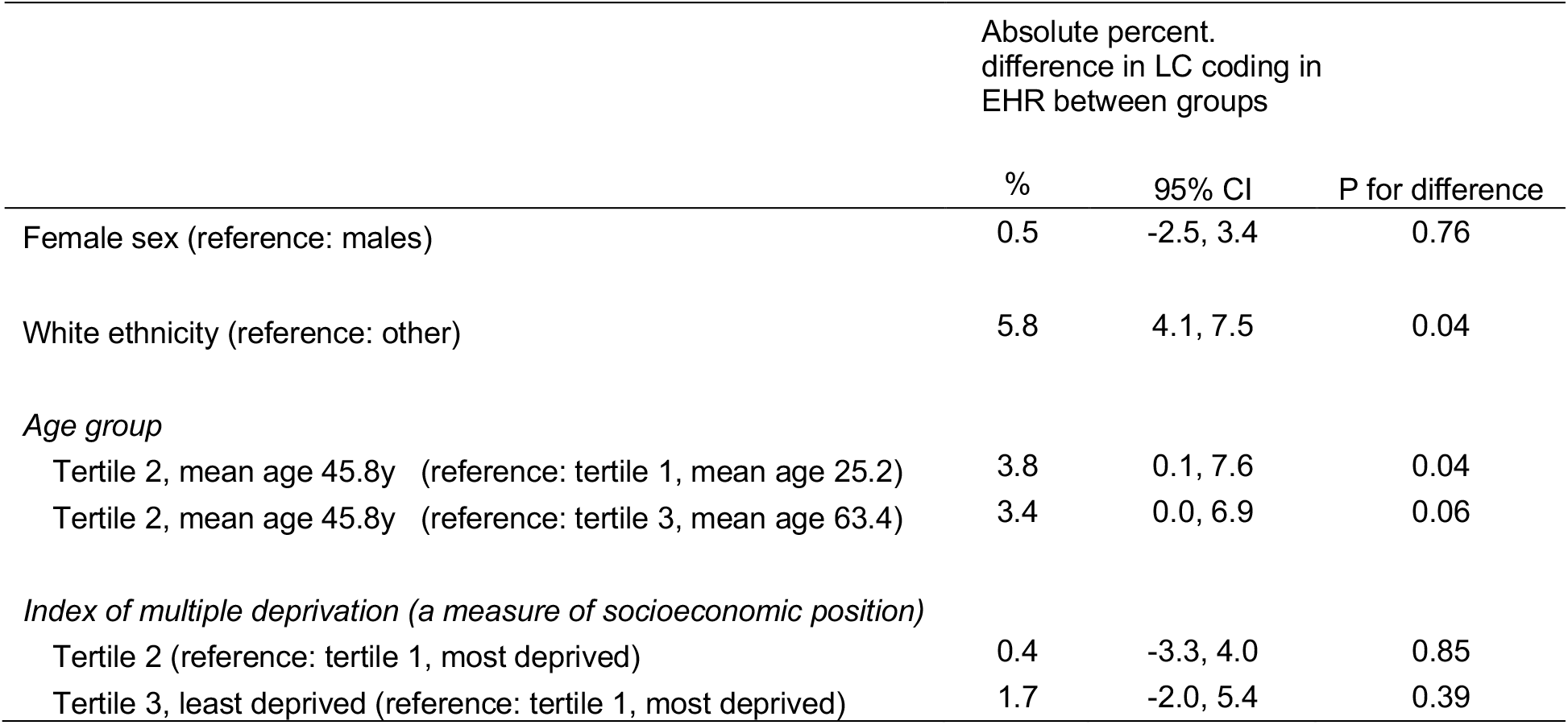
Percentage differences in LC coding in EHR between sociodemographic groups, among individuals with a history of self-reported LC (n ≤ 898)

|  | Absolute percent.<br>difference in LC coding in<br>EHR between groups |  |  |
| --- | --- | --- | --- |
|  | % | 95% CI | P for difference |
| Female sex (reference: males) | 0.5 | -2.5, 3.4 | 0.76 |
| White ethnicity (reference: other) | 5.8 | 4.1, 7.5 | 0.04 |
| *Age group* |  |  |  |
| Tertile 2, mean age 45.8y (reference: tertile 1, mean age 25.2) | 3.8 | 0.1, 7.6 | 0.04 |
| Tertile 2, mean age 45.8y (reference: tertile 3, mean age 63.4) | 3.4 | 0.0, 6.9 | 0.06 |
| *Index of multiple deprivation (a measure of socioeconomic position)* |  |  |  |
| Tertile 2 (reference: tertile 1, most deprived) | 0.4 | -3.3, 4.0 | 0.85 |
| Tertile 3, least deprived (reference: tertile 1, most deprived) | 1.7 | -2.0, 5.4 | 0.39 |

## Discussion

Caveats to these results should be considered. There are several reasons why self-reported symptoms may not lead to coding in EHRs. First, for self-reported symptoms to be apparent in EHRs they must be perceived by the sufferers as severe enough to warrant seeking care, where care is seen by sufferers as being potentially helpful. Second, where help is sought, data recording in clinical encounters is influenced by practitioner recognition of signs and symptoms, beliefs on the importance of these and appropriate coding in EHRs. This issue has been raised previously, with LC coding differing substantially in EHRs according to the clinical software system used to record them to an extent not plausibly explained by true differences in illness prevalence. ^3^ Other issues may have influenced our findings, including limits to EHR coverage (e.g. where LPS participants had emigrated from England), the possibility of clinical recording of LC existing in free text records that were inaccessible to us, and selection bias resulting from LPS participation. However, these considerations appear unlikely to provide a complete explanation for the substantial discrepancy we observed between perceived and reported LC and LC coding apparent in EHRs.

In conclusion, our data show a striking discrepancy between LC as perceived and reported by participants in LPS and evidence of LC recorded in their EHRs. This could reflect substantial unmet clinical need, in keeping with patient reports of difficulties accessing healthcare and sub-optimal recognition of, and response to, their illness when they do. ^4^ Our data also suggest that unmet needs may be higher amongst individuals of non-white ethnicity and possibly amongst the socially disadvantaged. Moreover, these results indicate potential shortcomings of epidemiological research on emerging conditions such as LC based on EHR- or LPS-based ascertainment alone, which may not be recognised or acknowledged in published research studies. These resources each have distinct strengths and weaknesses for identifying LC in populations, and this research illustrates the value of triangulation between LPS and EHR data where these are available in the same individuals and made accessible efficiently and acceptably through research resources such as the UK LLC.

## Data Availability

All data used in the present study are available to request from the UK Longitudinal Linkage Collaboration - the national Trusted Research Environment for UK longitudindal population studies (https://ukllc.ac.uk). The syntax used in this study is available from the UK LLC GitHub (https://github.com/UKLLC/LLC_0006).

https://github.com/UKLLC/LLC_0006

## Acknowledgements

This research was funded by the National Institute for Health and Care Research (NIHR; grants COV-LT-0009; MC_PC_20051). The UK LLC is an initiative of the UKRI-funded Longitudinal Health and Wellbeing National Core Study led by University College London and University of Bristol (Grant codes: MC_PC_20030, MC_PC_20059). JM is partly funded by the NIHR Applied Research Collaboration West. A full list of acknowledgments is provided in the supplementary materials. The authors declare no conflicts of interest.

