## Supplementary Materials for "The long COVID evidence gap: comparing self-reporting and clinical coding of long COVID using longitudinal study data linked to healthcare records"

### Supplementary material

### *Supplemental text*

#### *Study overview*

To define self-reported history of long COVID, we used data from the following eight UK-based longitudinal population-based studies (LPS; table S1), all of which implemented COVID-19-related web and postal questionnaires throughout the pandemic:

- The Avon Longitudinal Study of Parents and Children (ALSPAC) [1,2] – stratified here into a parent / partner sample (Generation zero; G0) and offspring sample (Generation 1; G1)
- 1970 British Cohort Study (BCS70)
- Medical Research Council (MRC) National Survey of Health and Development (NSHD) [3]
- Millennium Cohort Study (MCS) – stratified into an offspring and parent sample [4,5]
- National Child Development Study (NCDS)
- Next Steps
- TwinsUK[6]
- The UK Household Longitudinal Study (Understanding Society).

#### *COVID-19 and long COVID ascertainment from surveys*

COVID-19 status was defined by self-report, including reporting of test-confirmed and healthcare professional diagnosed disease. Long COVID was defined based on questions regarding symptom duration since acute illness, asked of individuals having reported COVID-19. All studies asked respondents to define symptom duration categorically (0-4 weeks, 4-12 weeks, or 12 or more weeks) with the exception of Understanding Society, which asked respondents to state numerically the number of weeks that symptoms persisted for. From these, a binary variable indicating long COVID – symptoms attributable to COVID-19 lasting 4+ weeks since acute disease -- was derived for each study, in accordance with NICE guidance. A full description of questionnaire items per study has been published previously.[7]

#### *Identifying long COVID codes among EHR using the UK LLC*

The UK Longitudinal Linkage Collaboration (UK LLC) is a Trusted Research Environment developed and operated by the Universities of Bristol and Edinburgh using an underlying 'Secure eResearch Platform' infrastructure (https://serp.ac.uk/) provided by Swansea University for longitudinal research. The UK LLC TRE is designed to host de-identified data from many interdisciplinary LPS; to systematically link these to participants' health, administrative and environmental records; and to provide a secure analysis environment. This project has been approved by UK LLC and its contributing data owners and information on this project and its outputs can be accessed via UK LLC's website (https://ukllc.ac.uk/data-use-register/). The UK LLC has ethical approval from the Health Research Authority Research Ethics Committee (Haydock Committee; ref: 20/NW/0446).

A list of designated COVID-19- and long COVID-related ICD-10 and SNOMED-CT codes (Table S2) was extracted from NHS Digital-issued hospital episode statistics (outpatient, inpatient, and accident and emergency records) and primary care records in the GDPPR for study participants with consent for EHR linkage. Dated codes were provisioned within the UK LLC for merging with data collected by LPS.

#### *Data on sociodemographic characteristics*

Data on age at self-reported long COVID were derived from dates of birth and the dates of survey completion in LPS.

Sex, ethnicity and index of multiple deprivation (IMD) were extracted from data provisioned by NHS Digital, and cross-checked against equivalent LPS values where available. Ethnicity in NHS Digital records were available with 2001 national category codes (Table S3) and these values collapsed into 'white' and 'other' categories, given the small sample sizes after cross-tabulations in the analyses. IMD was based on 2019 values for England representing relative deprivation at a small local area level, derived from local information on averaged measures of: i) income; ii) employment; iii) education, skills and training; iv) health and disability; v) crime; vi) barriers to housing and services; vii) living environment. All small areas in England are ranked from 1 (most deprived) to 32,844 (least deprived) based on a composite score of the seven measures of deprivation. Tertiles of IMD scores were derived for these analyses.

#### *Analyses*

Data from LPS were pooled for descriptive statistics. We estimated proportions (presented as percentages) with 95% confidence intervals calculated by the Agresti-Coull method.[8]

Tests of differences in proportions were two-sided in all instances with the exception of the use of a one-sided test for differences in long COVID coding by ethnicity (where we hypothesised individuals of white ethnicity were more likely to be coded than others). Univariable logistic regression was used to test for a trend in the likelihood of long COVID coding with increasing tertiles of IMD, with a one-sided significance test reflecting the hypothesis that individuals of higher socioeconomic position would be more likely to have received a code.

#### *Data and code availability*

Data used in this research are made available via the UK LLC, and cannot be used or shared outside its Trusted Research Environment. Researchers can apply to use UK LLC's resource using the procedure outlined in the UK LLC Data Access and Acceptable Use Policy. The UK LLC uses a system of managed open access for researchers who demonstrate their project is intended to improve the public good.

Code used to analyse longitudinal study data and linked EHR within the UK LLC are available at: https://github.com/UKLLC/LLC_0006.

#### *Extended acknowledgements*

*This research was funded by the National Institute for Health and Care Research (NIHR; grants COV-LT-0009; MC_PC_20051). The UK LLC is an initiative of the UKRI-funded Longitudinal Health and Wellbeing National Core Study led by University College London and University of Bristol (Grant codes: MC_PC_20030, MC_PC_20059). JM is partly funded by the NIHR Applied Research Collaboration West. A full list of acknowledgments is provided in the supplementary materials. The authors declare no conflicts of interest.*

This work uses data accessed within UK LLC's Trusted Research Environment (TRE), hosted by the Secure eResearch Platform (SeRP UK). We thank the SeRP UK Team at Swansea University and NHS Digital Health and Care Wales for providing the TRE's infrastructure and support. This work uses data provided by participants of the contributing LPS within the UK LLC TRE, which have been collected through their longitudinal study or as part of their care and support and/or interactions with UK government services. We wish to recognise and thank the study participants and each contributing LPS team, including data managers, administrators and those collecting data. We thank the following LPS for contributing data that made this research possible:

- Avon Longitudinal Study of Parents and Children (ALSPAC)[1,2]
- 1970 British Cohort Study (BCS70)
- Medical Research Council (MRC) National Survey of Health and Development (NSHD)[3]
- Millennium Cohort Study (MCS)[4] [5]

- National Child Development Study (NCDS)
- Next Steps
- TwinsUK[6]
- The UK Household Longitudinal Study (Understanding Society).

We thank the NHS and particularly NHS Digital for their work in curating participants' health records and for making these available for public benefit research designed to improve health services.

Finally, we acknowledge the wider input of researchers in the CONVALESCENCE Study Collaborative (https://www.ucl.ac.uk/covid-19-longitudinal-health-wellbeing/convalescence-study-collaborative) towards data collection and the development of infrastructure and code lists that underpinned this project.

*Table S1: Information on the study sample **

| LPS | Sample sizes | Age at COVID-19 questionnaire, mean years (SD) | Female sex, n (%) | Non-white ethnicity, n (%) * | LC definition | Combined LC, n (%; 95% CI) † | Debilitating LC, n (%; 95% CI) † | LC of any severity, n (%; 95% CI) † | Months between LC questionnaire and latest EHR data, mean (min-max) |
|---|---|---|---|---|---|---|---|---|---|
| ALSPAC G0 | 52 | 57.7 (4.4) | 52 (100.0) | 0 (0.0) | Any severity | | | | 17.6 (16.1-18.1) |
| ALSPAC G1 | 516 | 28.9 (0.5) | 323 (62.6) | 11 (2.5) | Any severity | | | | 17.2 (15.5-17.9) |
| BCS70 | 570 | 50.9 (0.0) | 326 (57.2) | 22 (4.7) | Debilitating | | | | 15.5 (14.6-15.9) |
| MCS parents | 601 | 51.9 (5.3) | 421 (70.0) | 35 (8.0) | Debilitating | | | | 15.4 (14.4-15.9) |
| MCS offspring | 837 | 20.0 (0.3) | 516 (61.7) | 132 (22.2) | Debilitating | | | | 15.3 (14.4-15.9) |
| NCDS | 534 | 63.0 (0.1) | 282 (52.8) | 15 (3.7) | Debilitating | | | | 15.5 (14.6-15.9) |
| NEXT STEPS | 513 | 31.0 (0.3) | 299 (58.3) | 117 (25.8) | Debilitating | | | | 15.4 (14.4-15.9) |
| NSHD | 47 | 74.9 (0.1) | 21 (44.7) | 0 (0.0) | Debilitating | | | | 15.5 (14.5-15.9) |
| TWINS UK | 572 | 51.9 (16.3) | 504 (88.1) | 16 (3.3) | Debilitating | | | | 21.6 (18.2-22.3) |
| UKHLS | 2,170 | 50.5 (15.3) | 1300 (59.9) | 172 (9.7) | Any severity | | | | 9.7 (8.0-16.1) |
| TOTAL | 6,412 | 44.9 (16.73) | 4044 (63) | 520 (10.1) | | 898 (14.0; 13.2, 14.9) | 395 (10.8; 9.8, 11.8) | 503 (18.4; 16.9, 19.9) | 14.2 (8.0-22.3) |

* LPS participants with linked EHR data whom self-reported COVID-19 in questionnaires in 2020-21

† Proportions with self-reported long COVID per individual LPS are not displayed to avoid low cell counts in some places

### *Table S2 Clinical codes for long COVID diagnosis and referral*

| Type | Code | Term |
|---|---|---|
| SNOMED-CT | 1119304009 | Chronic post-COVID-19 syndrome (disorder) |
| | 1326351000000108 | Post-COVID-19 syndrome resolved (finding) |
| | 1325021000000106 | Signposting to Your COVID Recovery |
| | 1325031000000108 | Referral to post-COVID assessment clinic |
| | 1325041000000104 | Referral to Your COVID Recovery rehabilitation platform |
| | 1325161000000102 | Post-COVID-19 syndrome |
| | 1325181000000106 | Ongoing symptomatic disease caused by severe acute respiratory syndrome coronavirus 2 |
| ICD-10 | U07.4 | Post COVID-19 condition |

### *Table S3 Ethnicity codes in EHR data*

| **Code** | **Description** |
|---|---|
| A | White - British |
| B | White - Irish |
| C | White - Any other White background |
| D | Mixed - White and Black Caribbean |
| E | Mixed - White and Black African |
| F | Mixed - White and Asian |
| G | Mixed - Any other mixed background |
| H | Asian or Asian British - Indian |
| J | Asian or Asian British - Pakistani |
| K | Asian or Asian British - Bangladeshi |
| L | Asian or Asian British - Any other Asian background |
| M | Black or Black British - Caribbean |
| N | Black or Black British - African |
| P | Black or Black British - Any other Black background |
| R | Other Ethnic Groups - Chinese |
| S | Other Ethnic Groups - Any other ethnic group |
| Z | Not stated |

Adapted from the NHS Data Model and Dictionary:

https://www.datadictionary.nhs.uk/attributes/ethnic_category_code_2001.html

### *Information on longitudinal population-based studies*

| ALSPAC: Avon Longitudinal Study of Parents and Children | |
|---|---|
| **Description of Study Population** (including citations and references if required) | Pregnant women resident in a defined area of the former county of Avon, UK with expected dates of delivery 1st April 1991 to 31st December 1992 were invited to take part in the study [1,2]. The initial number of pregnancies enrolled is 14,541 (14,676 foetuses), resulting in 14,062 live births and 13,988 children who were alive at 1 year of age. Further recruitment took place after the age of 7 years, the total sample size for analyses using any data collected after the age of 7 is therefore 15,454 pregnancies, resulting in 15,589 foetuses. Of these 14,901 were alive at 1 year of age.<br><br>[1]Boyd A, Golding J, Macleod J, Lawlor DA, Fraser A, Henderson J, Molloy L, Ness A, Ring S, Davey Smith G. Cohort Profile: The 'Children of the 90s'; the index offspring of The Avon Longitudinal Study of Parents and Children (ALSPAC). International Journal of Epidemiology 2013; 42: 111-127.<br>[2]Fraser A, Macdonald-Wallis C, Tilling K, Boyd A, Golding J, Davey Smith G, Henderson J, Macleod J, Molloy L, Ness A, Ring S, Nelson SM, Lawlor DA. Cohort Profile: The Avon Longitudinal Study of Parents and Children: ALSPAC mothers cohort. International Journal of Epidemiology 2013; 42:97-110. |
| **Acknowledgements** | We are extremely grateful to all the families who took part in this study, the midwives for their help in recruiting them, and the whole ALSPAC team, which includes interviewers, computer and laboratory technicians, clerical workers, research scientists, volunteers, managers, receptionists and nurses. |
| **Ethics** | Ethical approval for the study was obtained from the ALSPAC Law and Ethics committee and local research ethics committees (NHS Haydock REC: 10/H1010/70). |
| **Website** | http://www.bristol.ac.uk/alspac/researchers/access/ |

| BCS70: 1970 British Cohort Study | |
|---|---|
| **Description of Study Population** (including citations and references if required) | The 1970 British Cohort Study (BCS70) follows the lives of more than 17,000 people born in England, Scotland and Wales in a single week of 1970. Over the course of cohort members' lives, BCS70 has collected information on health, physical, educational and social development, and economic circumstances, among other factors.<br><br>Since the birth survey in 1970, there have been nine 'sweeps' of all cohort members at ages 5, 10, 16, 26, 30, 34, 38, 42 and most recently at 46 (a biomedical data collection). The Age 51 Sweep is currently in the field (2022).<br><br>Data have been collected from a number of different sources, including the midwife present at birth, parents of the cohort members, head and class teachers, school health service personnel and the cohort members themselves.<br><br>The data have been collected in a variety of ways, including via paper and electronic questionnaires, clinical records, medical examinations, biological samples, physical measurements, tests of ability, educational assessments and diaries.<br><br>The study is conducted by the Centre for Longitudinal Studies. |

| | |
|---|---|
| **Acknowledgements** | BCS70 is core-funded by the ESRC. |
| **Ethics** | Ethics approval has been obtained for each follow-up from an NHS Research Ethics Committee (REC) since 2000. In addition, separate REC approval is in place to cover the ongoing activities of the study in between major sweeps of data collection (i.e. Keeping in touch with and tracing cohort members; cleaning, documenting and providing access to the data for research; and linking data from administrative sources to survey data to increase the utility of the data for research). |
| **Website** | https://cls.ucl.ac.uk/cls-studies/bcs70/ |

| **MCS: Millennium Cohort Study** | |
|---|---|
| **Description of Study Population** (including citations and references if required) | The Millennium Cohort Study (MCS) is following the lives of young people born across England, Scotland, Wales and Northern Ireland in 2000-02. The study began with an original sample of 18,818 cohort members. The study is designed and led by the Centre for Longitudinal Studies (CLS) at University College London.<br><br>The broad aim of the study is to examine the impact that circumstances and experiences at one stage of life have on outcomes and achievements in later life. Since the baseline survey at age 9 months, there have been six major 'sweeps' at ages 3, 5, 7, 11, 14 and 17. The next sweep, at age 22, is currently under development.<br><br>Data have been collected from a number of different sources, including the cohort members and their parents and teachers. The data have been collected in a variety of ways, including via paper and electronic questionnaires, biological samples, physical measurements, tests of ability, and linked educational attainment and health records.<br><br>The information collected forms a high quality data resource for scientific investigations across a full range of domains of individuals' lives and across different points in time in them. The study has been designed to ensure comparability with other major cohort studies both in the UK and internationally and to permit the examination of links between social change and the changing experiences of different cohorts.<br><br>https://www.llcsjournal.org/index.php/llcs/article/view/410/0<br><br>https://academic.oup.com/ije/article/43/6/1719/703283 |
| **Acknowledgements** | MCS is core-funded by the ESRC and co-funded by a consortium of government departments. |
| **Ethics** | Ethics approval has been obtained for each follow-up from an NHS Research Ethics Committee (REC). In addition, separate REC approval is in place to cover the ongoing activities of the study in between major sweeps of data collection (i.e. Keeping in touch with and tracing cohort Members; cleaning, documenting and providing access to the data for research and linking data from administrative sources to survey data to increase the utility of the data for research. |
| **Website** | https://cls.ucl.ac.uk/cls-studies/mcs/ |

| **NCDS: National Child Development Study** |
|---|

| | |
|---|---|
| **Description of Study Population** (including citations and references if required) | The National Child Development Study (NCDS) is a continuing longitudinal study that seeks to follow the lives of all those living in Great Britain who were born in one particular week in 1958. Conducted by the Centre for Longitudinal Studies , the aim of the study is to improve understanding of the factors affecting human development over the whole lifespan. It collects information on physical and educational development, economic circumstances, employment, family life, health behaviour, wellbeing, social participation and attitudes.<br><br>The broad aim of the study is to examine the impact that circumstances and experiences at one stage of life have on outcomes and achievements in later life. Since the birth survey in 1958, there have been ten 'sweeps' of all cohort members at ages 7, 11, 16, 23, 33, 42, 44/5 (a biomedical collection) 46, 50 and most recently at 55. The Age 62 Sweep is currently in the field (2022).<br><br>Data have been collected from a number of different sources, including the midwife present at birth, parents of the cohort members, teachers, doctors and the cohort members themselves. The data have been collected in a variety of ways, including via paper and electronic questionnaires, clinical records, medical examinations, biological samples, physical measurements, tests of ability and educational assessments.<br><br>The information collected forms a high quality data resource for scientific investigations across a full range of domains of individuals' lives and across different points in time in them. The study has been designed to ensure comparability with other major cohort studies and to permit the examination of links between social change and the changing experiences of different cohorts.<br><br>https://cls.ucl.ac.uk/cls-studies/1958-national-child-development-study/ |
| **Acknowledgements** | NCDS is core-funded by the ESRC. |
| **Ethics** | Ethics approval has been obtained for each follow-up from an NHS Research Ethics Committee (REC) since 2000. In addition, separate REC approval is in place to cover the ongoing activities of the study in between major sweeps of data collection (i.e. Keeping in touch with and tracing cohort members; cleaning, documenting and providing access to the data for research; and linking data from administrative sources to survey data to increase the utility of the data for research). |
| **Website** | https://cls.ucl.ac.uk/cls-studies/ncds/ |

| **Next Steps** | |
|---|---|
| **Description of Study Population** (including citations and references if required) | Next Steps (previously known as the Longitudinal Study of Young People in England (LSYPE1)) is a major longitudinal study that follows the lives of around 16,000 people born in 1989-90. The first seven sweeps of the study (2004-2010) were funded and managed by the Department for Education and mainly focused on the educational and early labour market experiences of young people.<br><br>The study began in 2004 and included young people in Year 9 who attended state and independent schools in England. |

| | |
|---|---|
| | Following the initial survey at age 13-14, the cohort members were interviewed every year until 2010.<br><br>In 2013 the management of Next Steps was transferred to the Centre for Longitudinal Studies (CLS) at the IOE, UCL's Faculty of Education and Society. The first sweep conducted by CLS aimed to find out how the lives of the cohort members had turned out at age 25. It maintained the strong focus on education, but the content was broadened to become a more multi-disciplinary research resource.<br><br>The Age 32 Sweep is currently in the field (2022).<br><br>https://doc.ukdataservice.ac.uk/doc/5545/mrdoc/pdf/next_steps_userguide_to_the_redeposit_of_sweeps_1to7_may2020.pdf<br><br>https://doc.ukdataservice.ac.uk/doc/5545/mrdoc/pdf/nextsteps_age25_survey_user_guide_v3.pdf<br><br>https://cls.ucl.ac.uk/cls-studies/next-steps/ |
| **Acknowledgements** | Next Steps now is core-funded by the ESRC. |
| **Ethics** | Ethics approval is obtained for each follow-up from an NHS Research Ethics Committee (REC). In addition, separate REC approval is in place to cover the ongoing activities of the study in between major sweeps of data collection (i.e. keeping in touch with and tracing cohort members; cleaning, documenting and providing access to the data for research; and linking data from administrative sources to survey data to increase the utility of the data for research). |
| **Website** | https://cls.ucl.ac.uk/cls-studies/next-steps/ |

| **NSHD: Medical Research Council National Survey of Health and Development** | |
|---|---|
| **Description of Study Population (including citations and references if required)** | The MRC National Survey of Health and Development (NSHD) is a socially stratified birth cohort of 2,547 women and 2,815 men. It is a sample of all births in England, Scotland, and Wales that occurred in one week in March 1946, and consists of all single births to married women with a husband in non-manual and agricultural employment and 1 in 4 of all comparable births to women with a husband in manual employment.[1]<br><br>[1]Kuh et al. Cohort profile: updating the cohort profile for the MRC National Survey of Health and Development: a new clinic-based data collection for ageing research. Int J Epidemiol. 2011 Feb;40(1):e1-9. doi: 10.1093/ije/dyq231. |
| **Acknowledgements** | The UK Medical Research Council provides core funding for the MRC National Survey of Health and Development (MC_UU_00019/1). We are extremely grateful to the NSHD study members for their lifelong participation and continuing support; and to past and present members of the study teams, who helped to collect and process the data. |
| **Ethics** | Ethical approval for the study was obtained from the UK Research Ethics Committee (REC). |
| **Website** | https://skylark.ucl.ac.uk/ |

| **TwinsUK** |
|---|

| | |
|---|---|
| **Description of Study Population** (including citations and references if required) | TwinsUK is the largest adult twin registry in the UK and the most clinically detailed in the world. The national, population-based study was founded in 1992 and aims to investigate the genetic and environmental basis of a range of complex diseases and conditions. TwinsUK currently consists of over 15,700 volunteer adult twins (both monozygotic and dizygotic) who are between 18 to 104 years of age from around the UK (mean age 59)[1]. The cohort is predominantly female, and disease prevalence is broadly reflective of the UK population. Over 700,000 biological samples and extensive phenotypes have been collected longitudinally over 30 years.<br><br>[1]Verdi S, Abbasian G, Bowyer RCE, et al.: TwinsUK: The UK Adult Twin Registry Update. Twin Res Hum Genet. 2019; 22(6): 523–529. |
| **Acknowledgements** | We thank TwinsUK members for their participation and the TwinsUK operations team for coordinating and undertaking twin clinic visits and data and sample collections.<br><br>TwinsUK is funded by the Wellcome Trust, Medical Research Council, Versus Arthritis, European Union Horizon 2020, Chronic Disease Research Foundation (CDRF), Zoe Ltd, the National Institute for Health and Care Research (NIHR) Clinical Research Network (CRN) and Biomedical Research Centre based at Guy's and St Thomas' NHS Foundation Trust in partnership with King's College London. |
| **Ethics** | All collections of TwinsUK data have received ethical approval associated with TwinsUK Biobank (19/NW/0187), TwinsUK (EC04/015) or Healthy Ageing Twin Study (H.A.T.S) (07/H0802/84) from NHS Research Ethics Committees. Linkage to health and environmental records is also covered by approval from the Health Research Authority (19/CAG/0223). |
| **Website** | https://twinsuk.ac.uk/resources-for-researchers/access-our-data/ |

| **Understanding Society – the UK Household Longitudinal Study** | |
|---|---|
| **Description of Study Population** (including citations and references if required) | Understanding Society, the UK Household Longitudinal Study, is a longitudinal survey of the members of ~40,000 households (at Wave 1, 2009-10) in the United Kingdom. The survey sample consists of a large General Population Sample (~26,000 households) plus three other components: the Ethnic Minority Boost Sample (~4,000 households), the former British Household Panel Survey sample (~8,000 households) and the Immigrant and Ethnic Minority Boost Sample (~2,900 households, added at Wave 6). Household and individual interviews are conducted annually. The study is multi-topic and multi-purpose.<br><br>From April 2020 to September 2021, participants from the main Understanding Society sample were asked to complete nine short web-surveys (with a telephone option in some months). The COVID-19 study covered the changing impact of the pandemic on the welfare of UK individuals, families and wider communities. ~18,000 individuals provided a full or partial interview at Wave 1 (April 2020).<br><br>At Wave 8 of the COVID-19 study, 8477 participants provided consent to link their survey data to administrative health records. |

| | |
|---|---|
| **Acknowledgements** | Understanding Society is an initiative funded by the Economic and Social Research Council and various Government Departments, with scientific leadership by the Institute for Social and Economic Research, University of Essex, and survey delivery by NatCen Social Research and Kantar Public<br><br>The COVID-19 study (2020-2021) was funded by the Economic and Social Research Council and the Health Foundation. Serology testing was funded by the COVID-19 Longitudinal Health and Wealth – National Core Study. Fieldwork for the web survey was carried out by Ipsos MORI and for the telephone survey by Kantar. |
| **Ethics** | The University of Essex Ethics Committee has approved all data collection on Understanding Society main study, COVID-19 surveys and innovation panel waves, including asking consent for all data linkages except to health records.<br><br>Approval for asking consent for health record linkage and for the collection of blood and subsequent serology testing in the March 2021 wave of the COVID-19 study was obtained from London – City & East Research Ethics Committee (21/HRA/0644). |
| **Website** | https://ukllc.ac.uk or https://ukdataservice.ac.uk |
